# Large Language Model-Based Entity Extraction Reliably Classifies Pancreatic Cysts and Reveals Predictors of Malignancy: A Cross-Sectional and Retrospective Cohort Study

**DOI:** 10.1101/2025.07.15.25331413

**Authors:** Anthony Papale, Robert Flattau, Nandan Vithlani, Deepti Mahajan, Yonah Ziemba, Tiffany Zavadsky, Anthony Carvino, Daniel King, Sandeep Nadella

## Abstract

Pancreatic cystic lesions (PCLs) are often discovered incidentally on imaging and may progress to pancreatic ductal adenocarcinoma (PDAC). PCLs have a high incidence in the general population, and adherence to screening guidelines can be variable. With the advent of technologies that enable automated text classification, we sought to evaluate various natural language processing (NLP) tools including large language models (LLMs) for identifying and classifying PCLs from radiology reports. We correlated our classification of PCLs to clinical features to identify risk factors for a positive PDAC biopsy. We contrasted a previously described NLP classifier to LLMs for prospective identification of PCLs in radiology. We evaluated various LLMs for PCL classification into low-risk or high-risk categories based on published guidelines. We compared prompt-based PCL classification to specific entity-guided PCL classification. To this end, we developed tools to deidentify radiology and track patients longitudinally based on their radiology reports. Additionally, we used our newly developed tools to evaluate a retrospective database of patients who underwent pancreas biopsy to determine associated factors including those in their radiology reports and clinical features using multivariable logistic regression modelling. Of 14,574 prospective radiology reports, 665 (4.6%) described a pancreatic cyst, including 175 (1.2%) high-risk lesions. Our Entity-Extraction Large Language Model tool achieved recall 0.992 (95% confidence interval [CI], 0.985-0.998), precision 0.988 (0.979-0.996), and F1-score 0.990 (0.985-0.995) for detecting cysts; F1-scores were 0.993 (0.987-0.998) for low-risk and 0.977 (0.952-0.995) for high-risk classification. Among 4,285 biopsy patients, 330 had pancreatic cysts documented ≥6 months before biopsy. In the final multivariable model (AUC = 0.877), independent predictors of adenocarcinoma were change in duct caliber with upstream atrophy (adjusted odds ratio [AOR], 4.94; 95% CI, 1.30-18.79), mural nodules (AOR, 11.02; 1.81-67.26), older age (AOR, 1.10; 1.05-1.16), lower body mass index (AOR, 0.86; 0.76-0.96), and total bilirubin (AOR, 1.81; 1.18-2.77). Automated NLP-based analysis of radiology reports using LLM-driven entity extraction can accurately identify and risk-stratify PCLs and, when retrospectively applied, reveal factors predicting malignant progression. Widespread implementation may improve surveillance and enable earlier intervention.

## Introduction

Pancreatic cancer is among the leading causes of cancer-related death in the United States, with an estimated 66,440 new cases and 51,570 deaths in 2024.^1–3^ Pancreatic ductal adenocarcinoma (PDAC) accounts for approximately 90% of cases.^4^ Five-year survival remains dismal at about 10%, largely because most patients present at a late stage.^5^

PDAC can arise from precursor lesions, including pancreatic cystic lesions (PCLs), which are often discovered incidentally on imaging and present an opportunity for earlier intervention. PCLs have an estimated global prevalence between 2-18%, increasing with age.^6,7^ The majority are benign, yet certain mucinous subtypes, including intraductal papillary mucinous neoplasms (IPMNs) and mucinous cystic neoplasms (MCNs), carry significant malignant potential.^8,9^ The American Gastroenterological Association (AGA) and American College of Gastroenterology (ACG) guidelines recommend distinguishing low-risk cysts suitable for surveillance from high-risk cysts that may require endoscopic evaluation or resection.^10,11^ Despite these guidelines, real-world adherence varies, with many patients lacking appropriate surveillance or specialty referral.^12–14^

Recent advances in artificial intelligence (AI), particularly natural language processing (NLP), allow for automated interpretation of radiology reports to detect PCLs.^15^ Early rule-based NLP methods achieved high sensitivity, while modern deep learning, including transformer-based models, provide enhanced contextual understanding.^16,17^ Large language models (LLMs) have demonstrated expert-level interpretation in certain medical text tasks, although their performance in evaluating pancreatic imaging findings remains unclear.^18,19^

In this two-part study, we first evaluate and validate several NLP-based tools for identifying and classifying PCLs using existing computed tomography (CT) and magnetic resonance imaging (MRI) reports. We then apply these tools retrospectively to assess patients with biopsy-confirmed PDAC or nonmalignant findings, examining whether certain imaging features or clinical factors predict malignant transformation. We sought to demonstrate the potential of AI-driven radiology report analysis to improve surveillance and facilitate earlier intervention for PCLs.

## Methods

### Radiology Report Analysis and NLP Approaches

Data were collected under Northwell Health Institutional Review Board guidelines. We queried all CT and MRI reports from 15 hospitals in our health system between February 14 and March 6, 2024, for patients 18 years or older, yielding 14,574 free-text reports. Each report was manually reviewed to identify pancreatic abnormalities and PCLs. A cyst was considered high-risk if, when present, the recommended next step in management was endoscopic ultrasound (EUS) ± fine-needle aspiration (FNA) per ACG recommendations. High-risk cysts met at least one of the following criteria: main pancreatic duct diameter or dilation >5 mm; cyst size ≥3 cm; cyst size ≥2 cm in lesions not described as IPMN or MCN; presence of a mural nodule or solid component; or a change in main duct caliber with upstream pancreatic atrophy.

Several NLP techniques were applied to automate cyst identification and risk stratification. First, a previously described transformer-based model, iNav, was used to identify PCLs from radiology report text.^20^ iNav was derived from RadBERT and fine-tuned to detect abnormal pancreatic imaging findings.^21^ iNav assigns a single label to the report when an abnormality is present but does not perform risk stratification. We developed a PHI-compliant application that interfaces with the Northwell AI Hub API, which hosts multiple large-language models. The application securely sends each radiology report to the AI Hub, where the model parses the text according to predefined instructions.

Two versions of the LLM tool were implemented. In Open-Prompt LLM (opLLM), the model received broad, free-text questions asking whether a cyst was present and, if so, whether it was high risk. We tested three prompt variants (Supplemental Table 1). In Entity-Extraction LLM (eeLLM), the model was prompted to determine whether a PCL was present and to extract specific features from the text, including cyst size and main duct diameter (Supplemental Figure 1). These extracted entities were then processed by an algorithm to risk-stratify each lesion. We compared iNav, opLLM Variant 1, and eeLLM for cyst identification, then tested all three opLLM variants against eeLLM for risk stratification. The prompts for opLLM and eeLLM were initially developed using the generative pretrained transformer 4 (GPT-4) LLM. We subsequently compared multiple LLMs (Gemini 1.0, Gemini 1.5 Flash, Gemini 1.5 Pro, PaLM 2, GPT-4o, and GPT-4) using the best-performing LLM tool.

### Retrospective Evaluation of PCLs and Subsequent PDAC Risk

We retrospectively analyzed all patients 18 years or older who underwent pancreatic biopsy at the same 15 hospitals between January 2019 and November 2024. Biopsies confirming PDAC were deemed positive. For multiple biopsies, we used the earliest positive or most recent negative date. Patients without prior CT or MRI of the abdomen or chest, or whose first imaging occurred within six months of the biopsy, were excluded. For the remaining patients, all radiology reports predating the biopsy date (August 2016 to November 2024) were manually reviewed and parsed with eeLLM. Patients whose imaging showed only normal pancreatic findings or solid pancreatic masses, or whose cyst was first identified within six months of biopsy, were excluded to allow sufficient time for potential cyst progression. The final cohort consisted solely of patients with a PCL, classified as high-risk or low-risk according to the criteria described above. We separated biopsy-positive and biopsy-negative patients to compare demographics, imaging features, and laboratory values.

### Statistical Analysis

Statistical analyses were conducted in Python version 3.11.5. Analyses with bilateral *P* < 0.05 were considered statistically significant. Performance was evaluated using recall, precision, and F1-score. The 95% confidence intervals (CIs) were computed with bootstrapping (10,000 samples) using SciPy 1.13. Demographics were evaluated using chi-square (sex, race) or Mann-Whitney tests (age, body mass index [BMI]). Univariable logistic regression was used to calculate odds ratios (ORs) with 95% CI for imaging features. Serum labs were analyzed via logistic regression and Mann-Whitney tests, using the time closest to cyst identification (up to one year before or one month after). Continuous laboratory variables with absolute skewness greater than 1 were log-transformed using the natural logarithm. These analyses were repeated in both high-risk and low-risk subgroups. Finally, three multivariable logistic regression models were created: (1) imaging features only, (2) imaging features and demographics, and (3) imaging features, demographics, and labs. Adjusted odds ratios (AOR) with 95% CI were calculated. Model discrimination was evaluated with area under the curve (AUC) via receiver operating characteristic (ROC) curves and calibration with the Hosmer-Lemeshow (H-L) statistic.

## Results

### Radiology Report Analysis and NLP Approaches

#### Prevalence of Pancreatic Cysts in Prospective Radiology Reports

A total of 14,574 prospectively obtained free-text radiology reports from 13,410 patients were manually reviewed, of which 2,301 (15.8%) contained a pancreatic abnormality (Supplemental Table 2). Among these, 665 (4.6%) reports described a pancreatic cyst, with 175 (1.2%) high-risk and 490 (3.4%) low-risk cysts. Overall, 612 (4.6%) of the 13,410 unique patients had a cyst, including 159 (1.2%) high-risk PCLs.

#### eeLLM Demonstrates Superior Cyst Identification

For cyst detection (Figure 1a), eeLLM achieved highest performance with recall 0.992 (95% CI, 0.985-0.998), precision 0.988 (0.979-0.996), and F1-score 0.990 (0.985-0.995). opLLM Variant 1 performed similarly with recall 0.991 (95% CI, 0.983-0.997), precision 0.972 (0.959-0.984), and F1-score 0.981 (0.974-0.988). iNav showed substantially lower performance with recall 0.857 (95% CI, 0.830-0.883), precision 0.551 (0.520-0.581), and F1-score 0.671 (0.644-0.695).

**Figure 1.**
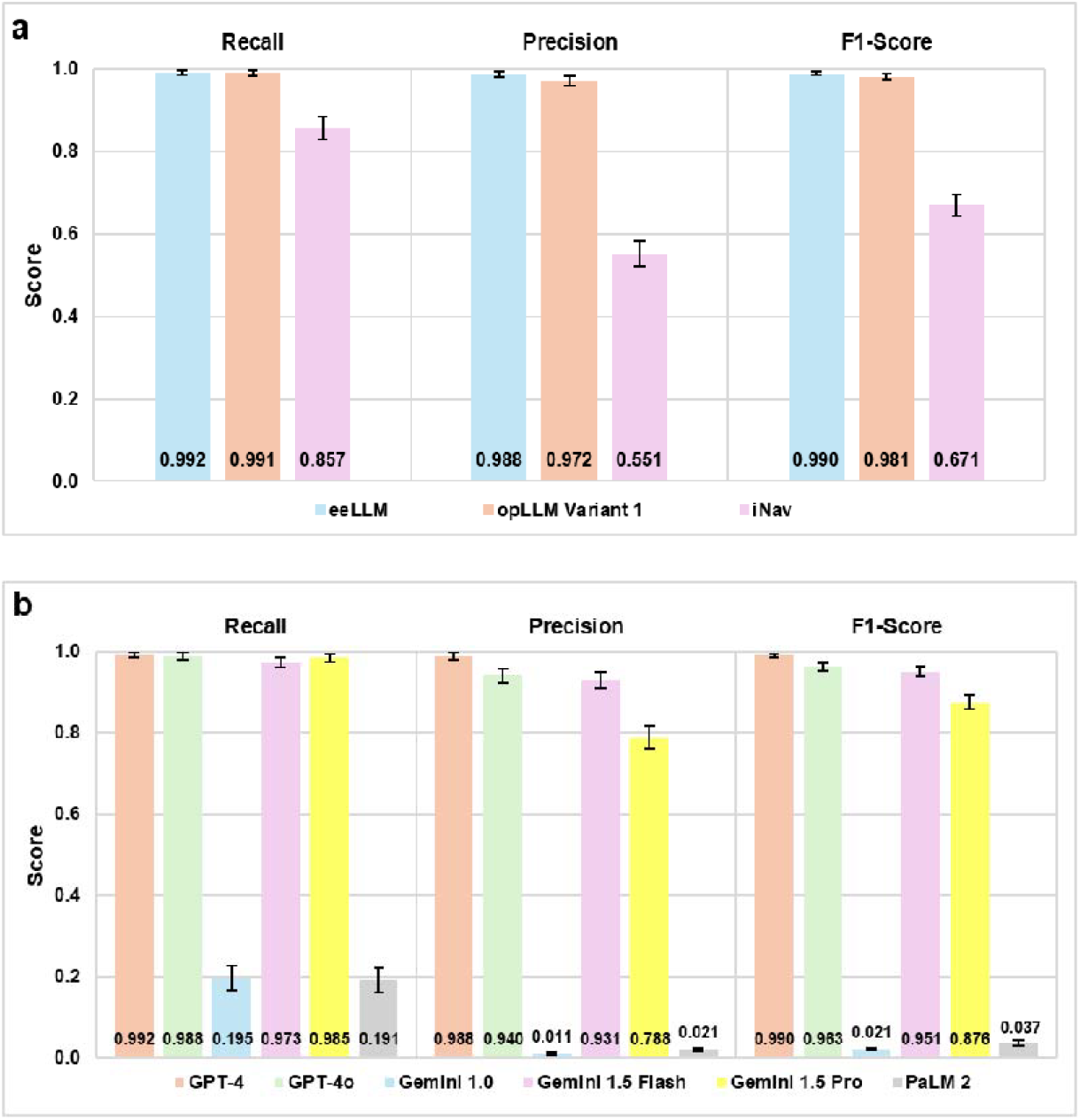
Performance Metrics of Automated Tools for Pancreatic Cyst Identification from Free-Text Radiology Reports. Bar graphs display recall, precision, and F1-scores with 95% CIs for (A) three NLP-based tools identifying pancreatic cysts (n=665) from free-text radiology reports (n=14,574) and (B) six LLMs performing cyst identification via the eeLLM tool. *Abbreviations:* CI, confidence intervals; eeLLM, Entity-Extraction Large Language Model; NLP, natural language processing; opLLM, Open-Prompt Large Language Model.

#### GPT-4 Outperforms Other LLMs in Cyst Identification

When comparing the six LLMs using eeLLM for cyst identification (Figure 1b), GPT-4 was the strongest performing model with recall 0.992 (95% CI, 0.985-0.998), precision 0.988 (0.979-0.996), and F1-score 0.990 (0.985-0.995). GPT-4o and Gemini 1.5 Pro performed similarly, with recalls 0.988 (95% CI, 0.979-0.996) and 0.985 (0.975-0.994), whereas Gemini 1.0 and PaLM 2 showed lower recalls of 0.195 (0.165-0.226) and 0.191 (0.161-0.221).

#### eeLLM Outperforms opLLM for Pancreas Cyst Classification

Our eeLLM tool outperformed all opLLM variants in classifying low-risk and high-risk cysts (Figure 2a and 2b). For low-risk lesions, the eeLLM tool achieved highest recall 0.996 (95% CI, 0.989-1.000), precision 0.990 (0.980-0.998), and F1-score 0.993 (0.987-0.998); opLLM variants had recalls from 0.955 to 0.976 and lower F1-scores. A similar pattern appeared for high-risk classification, with eeLLM reaching recall 0.977 (95% CI, 0.952-0.995), precision 0.977 (0.953-0.995), and F1-score 0.977 (0.960-0.992), whereas opLLM variants ranged from recalls 0.171 to 0.389.

**Figure 2.**
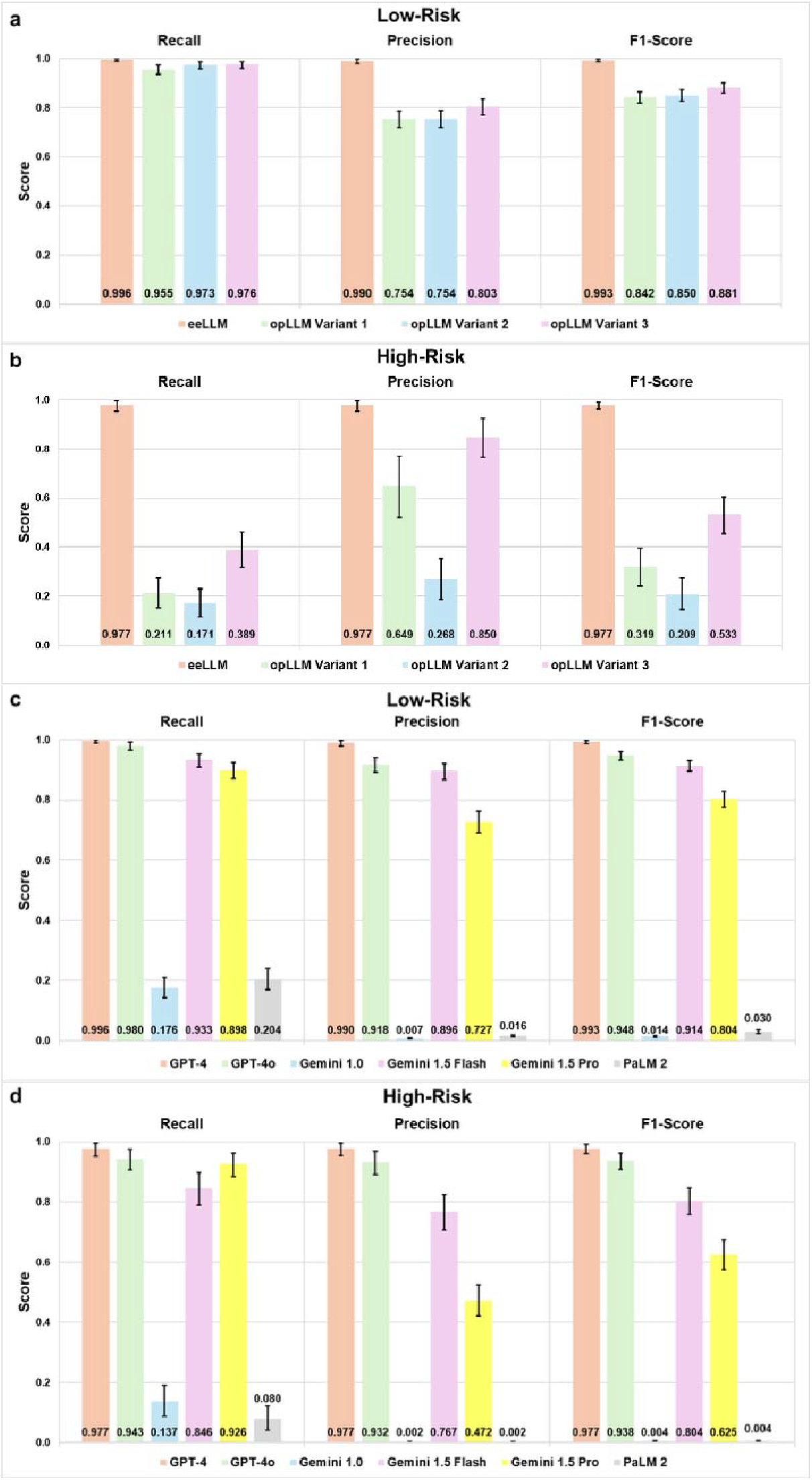
Performance Metrics of Automated Tools for Pancreatic Cyst Risk Stratification from Free-Text Radiology Reports. Bar graphs display recall, precision, and F1-scores with 95% CIs for (a) low-risk cyst (n=490) and (b) high-risk cyst (n=175) stratification from free-text radiology reports (n=14,574) by the eeLLM tool and three opLLM variants, and for (c) low-risk cyst and (d) high-risk cyst stratification by six LLMs via the eeLLM tool. *Abbreviations:* CIs, confidence interval; eeLLM, Entity-Extraction Large Language Model; NLP, natural language processing; opLLM, Open-Prompt Large Language Model.

#### GPT-4 Outperforms Other LLMs in Cyst Classification

For low-risk classification (Figure 2c), GPT-4 again attained highest recall 0.996 (95% CI, 0.989-1.000), precision 0.990 (0.980-0.998), and F1-score 0.993 (0.987-0.998), followed by GPT-4o with recall 0.980 (0.966-0.992), precision 0.918 (0.893-0.940), and F1-score 0.948 (0.933-0.961). Gemini 1.0 and PaLM 2 had much lower recalls of 0.176 (95% CI, 0.143-0.209) and 0.204 (0.169-0.240), respectively. For high-risk cyst classification (Figure 2d), GPT-4 achieved highest recall 0.977 (95% CI, 0.952-0.995), precision 0.977 (0.953-0.995), and F1-score 0.977 (0.960-0.992), followed by GPT-4o with recall 0.943 (0.907-0.975), precision 0.932 (0.892-0.968), and F1-score 0.938 (0.909-0.962). Gemini 1.0 and PaLM 2 performed poorly with recalls 0.137 (95% CI, 0.087-0.190) and 0.080 (0.041-0.122), respectively.

### eeLLM Tool Identifies BMI and Age as Risk Factors for a Positive PDAC Biopsy in Patients with preexisting PCLs

#### Cohort Characteristics

Between January 2019 and November 2024, 4,285 patients in our health system underwent pancreatic biopsy. After excluding those without prior imaging or whose first imaging study occurred within six months of the most recent biopsy, 1,059 remained. Of these, 411 had pancreatic cysts (275 high-risk, 136 low-risk), whereas 648 were excluded for having only normal pancreatic features or non-cystic abnormalities, or a concurrent solid pancreatic mass. An additional 81 patients were excluded because their cyst was first documented within six months of biopsy, yielding a final cohort of 330. Among these 330, 225 were high-risk (25 PDAC-positive, 200 PDAC-negative), and 105 were low-risk (10 PDAC-positive, 95 PDAC-negative). Figure 3 presents a flow diagram of these steps. We assessed our eeLLM tool on the cohort of 1,059 patients, yielding similar performance to the first cohort with recall 0.990 (95% CI, 0.980-0.998), precision 0.991 (0.983-1.000), and F1-score 0.991 (0.984-0.997) for cyst identification, recall 0.956 (0.918-0.986), precision 0.971 (0.938-0.993), and F1-score 0.963 (0.938-0.984) for low-risk classification, and recall 0.978 (0.960-0.993), precision 0.979 (0.959-0.993), and F1-score 0.979 (0.965-0.989) for high-risk classification.

**Figure 3.**
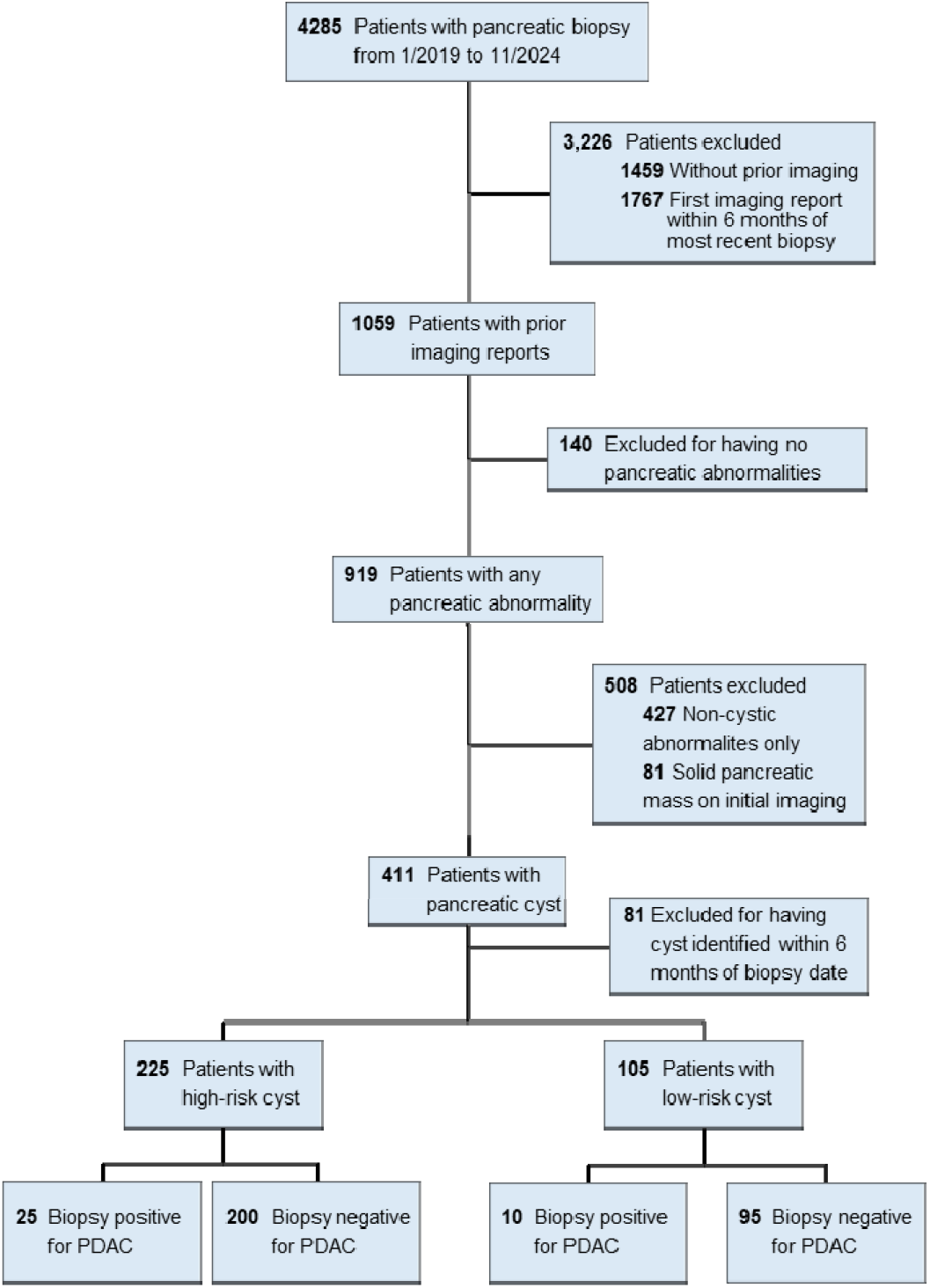
Retrospective Study Flow Diagram. *Abbreviations:* CT, computed tomography; MR, magnetic resonance; PDAC, pancreatic ductal adenocarcinoma.

#### Evaluation of risk factors associated with a positive biopsy in PCLs

Table 1 indicates that sex did not differ by PDAC status in any group, and race was also nonsignificant (Supplemental Table 3 for race distribution). Age was consistently higher in PDAC-positive patients, with median ages of 78 vs 71 years in the overall cyst and low-risk cohorts (*P*<0.001 and *P*=0.044), and 77 vs 70 years (*P*<0.001) in the high-risk subgroup. Median BMI was significantly lower in PDAC-positive patients in the entire cyst cohort (24.7 vs 28.2, *P*<0.001) and high-risk subgroup (24.7 vs 28.5, *P*<0.001) but not in the low-risk group.

**Table 1.** Demographic characteristics for PDAC-positive versus PDAC-negative patients. Sex and Race were evaluated using chi-square tests; Age and BMI were evaluated using the Mann-Whitney U test. Significant differences (*P*<0.05) are marked with an asterisk (*). *Abbreviations*: BMI, body mass index; IQR, interquartile range; PDAC, pancreatic ductal adenocarcinoma.

|  | All Cysts |  |  | High-Risk Cysts |  |  | Low-Risk Cysts |  |  |
| --- | --- | --- | --- | --- | --- | --- | --- | --- | --- |
|  | Positive Biopsy (n=35) | Negative Biopsy (n=295) | P-value | Positive Biopsy (n=25) | Negative Biopsy (n=200) | P-value | Positive Biopsy (n=10) | Negative Biopsy (n=95) | P-value |
| **Sex** | Female=19; Male=16 | Female=161; Male=134 | 1.000 | Female=13; Male=12 | Female=101; Male=99 | 1.000 | Female=6; Male=4 | Female=60; Male=35 | 1.000 |
| **Median Age in Years (IQR)** | 78.0 (13.0) | 71.0 (18.5) | <0.001* | 77.0 (14.0) | 70.0 (19.0) | <0.001* | 78.0 (8.5) | 71.0 (14.0) | 0.044* |
| **Race** | $\chi^2 = 4.74$ | | 0.449 | $\chi^2 = 4.89$ | | 0.430 | $\chi^2 = 2.97$ | | 0.560 |
| **Median BMI (IQR)** | 24.7 (5.9) | 28.2 (7.9) | <0.001* | 24.7 (4.4) | 28.5 (8.4) | <0.001* | 24.5 (10.6) | 27.7 (7.3) | 0.292 |

Supplemental Table 4 shows results of univariable logistic regression of seven imaging features (Supplemental Table 5 for exact counts). Because cyst size, main duct dilation, duct caliber change with atrophy, and mural nodules define a high-risk cyst, these variables were not separately analyzed in low-risk lesions. Duct dilation was observed in 45.7% of PDAC-positive vs 18.6% of PDAC-negative patients (OR 3.67; 95% CI 1.78-7.60; *P*=0.001) overall, and 64.0% vs 27.5% (4.69; 1.96-11.23; *P*<0.001) in high-risk cysts. Duct caliber change with atrophy appeared in 25.7% vs 8.1% (OR 3.91; 95% CI 1.65-9.29; *P*=0.004) overall, and 36.0% vs 12.0% (4.12; 1.64-10.36; *P*=0.004) in high-risk cysts. Mural nodules were found in 14.3% vs 3.4% (OR 4.75; 95% CI 1.52-14.82; *P*=0.014) overall, rising to 20.0% vs 5.0% (4.75; 1.48-15.28; *P*=0.016) in high-risk cysts. Although cyst size was nonsignificant (OR 0.51; 95% CI 0.25-1.03; *P*=0.070) overall, it reached significance (0.20; 0.08-0.51; *P*=0.001) in high-risk PCLs. Enhancing cyst wall, septations, and pancreatitis were not significant.

Among log-transformed laboratory values (Supplemental Table 6), only carbohydrate antigen (CA) 19-9 was significantly associated with a diagnosis of PDAC in the entire cohort (OR 3.42; 95% CI 1.24-9.45; *P*=0.018) and high-risk subgroup (OR 3.20; 1.18-8.67; *P*=0.023). Mann-Whitney comparisons (Supplemental Table 7) of CA 19-9 values were also higher in PDAC-positive patients (70.85 vs 12.0 U/mL; *P*=0.006 overall; 70.85 vs 12.0 U/mL; *P*=0.009 high-risk subgroup). In the low-risk subgroup, CA 19-9, carcinoembryonic antigen (CEA), and alpha-fetoprotein (AFP) lacked sufficient observations for analysis. No other laboratory parameters were significant. Laboratory study counts are provided in Supplemental Table 7.

#### Multi-modal integration of pathology, radiology reports and laboratory information reveals BMI and age as associated risk factors for a positive biopsy in PCLs

In Model A (n=330, imaging features only), main duct diameter (AOR 2.65; 95% CI 1.13-6.19; *P*=0.024), mural nodules (4.74; 1.40-16.08; *P*=0.013), and cyst size (0.41; 0.19-0.88; *P*=0.021) were significant, while duct caliber change trended toward significance. Model B (n=330, imaging and demographics) confirmed age (AOR 1.08; 95% CI 1.04-1.12; *P*<0.001), BMI (0.91; 0.83-0.98; *P*=0.017), main duct diameter (2.60; 1.04-6.53; *P*=0.042), mural nodules (5.21; 1.29-20.98; *P*=0.020), and cyst size (0.42; 0.19-0.94; *P*=0.036) as independent predictors, while duct caliber change was not significant. In Model C (n=261, imaging, demographics, and labs), CA 19-9 was excluded due to near-complete separation. Total bilirubin (AOR 1.81; 95% CI 1.18-2.77; *P*=0.006) became significant, while main duct diameter (1.52; 0.44-5.30; *P*=0.512) and cyst size (0.36; 0.13-1.01; *P*=0.052) lost significance. Change in duct caliber with atrophy emerged as an independent factor (AOR 4.94; 95% CI 1.30-18.79; *P*=0.019), along with mural nodules (11.02; 1.81-67.26; *P*=0.009), age (1.10; 1.05-1.16; *P*<0.001), and BMI (0.86; 0.76-0.96; *P*=0.008). We observed a moderate correlation between main duct diameter and duct caliber change with atrophy (r=0.38), which may explain why main duct diameter lost significance when additional covariates were introduced. Figure 4 shows a forest plot with AOR and 95% CI of each model. ROC curves in Supplemental Figure 2 demonstrate Model C had the highest AUC of 0.877 versus the lowest AUC of 0.721 for Model A, while calibration plots in Supplemental Figure 3 demonstrated acceptable calibration across all models with HL *P*≥0.665.

**Figure 4.**
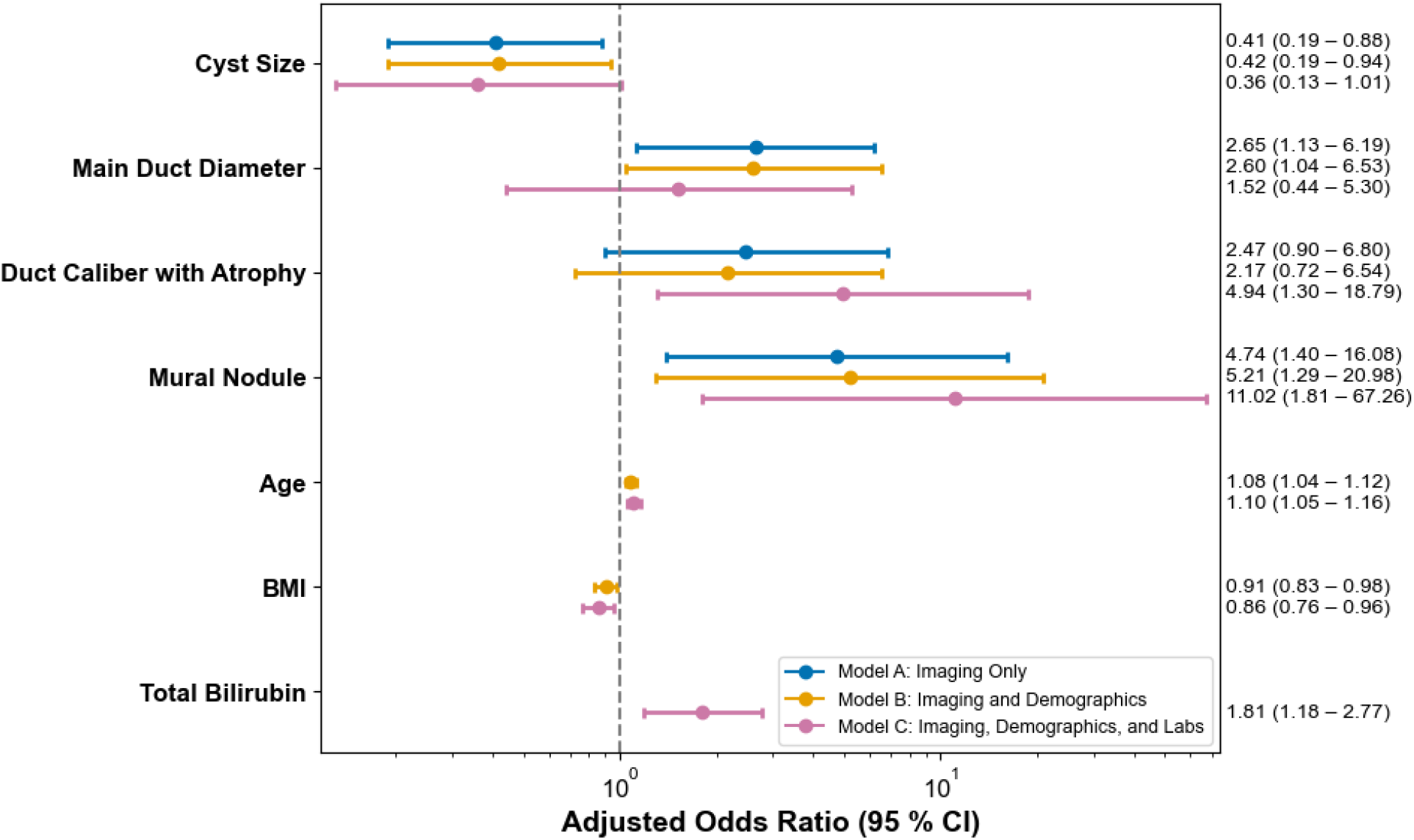
Forest plot containing adjusted OR with 95% CI for three multivariable logistic regression models. *Abbreviations:* BMI, body mass index.

## Discussion

We leveraged NLP-based tools to identify and risk-stratify PCLs from free-text radiology reports, then applied these tools to assess factors predicting malignant progression. Two prior studies investigated using automated NLP to detect PCLs: one employed rule-based expressions while another deployed a transformer-based model.^16,22^ Our study expands on this by being among the first to investigate using LLMs to perform cyst identification and risk stratification, evaluate how variations in prompting strategies can influence LLM performance of these tasks, and integrate both tasks into a single pipeline.

In our initial analysis, both the eeLLM and opLLM tools outperformed the transformer-based iNav for cyst identification, likely reflecting the superior language understanding of LLMs. eeLLM consistently achieved performance near manual review for cyst detection and risk classification, whereas the best opLLM variant performed significantly worse in classification. These results suggest that prompting LLMs to extract discrete features from text is superior to broader, open-ended instructions. When tested across multiple LLMs, GPT-4 and GPT-4o yielded higher recall and F1-scores, likely reflecting their larger parameter sets. Notably, GPT-4 outperformed GPT-4o, suggesting that newer versions do not always guarantee superior performance if prompts are not tailored to them, and as such prompt design should be reevaluated whenever adopting a new LLM.

For our second analysis, an initial cohort of 4,285 patients who underwent pancreatic biopsy was narrowed to 330 individuals who had pancreatic cysts identified on imaging at least six months before biopsy. Of these 330, 25 of 225 high-risk cysts (11.1%) and 10 of 105 low-risk cysts (9.5%) were positive for PDAC, highlighting the need to better understand the reasons for which low-risk lesions progress. Univariable analysis showed that older age and lower BMI were associated with PDAC across the overall cohort, findings that persisted in the final multivariable models. Age correlates with increasing PCL prevalence and malignant transformation, with studies reporting higher mean age in malignancy (67 vs 62 years, *P*<0.05) and a hazard ratio of 1.03 per year (95% CI, 1.00-1.06).^23,24^ Conversely, higher BMI has been correlated with increased malignancy risk, though it should be noted that these studies specifically assessed patients with IPMNs or MCNs.^25,26^

CA 19-9 was the only laboratory parameter associated with PDAC in univariable analysis, with significantly higher values in PDAC-positive patients. These results align with two meta-analyses that demonstrated CA 19-9 as a predictor of malignancy in pancreatic cystic neoplasms.^27,28^ Although CA 19-9 was excluded from our final multivariable model due to near-complete separation, total bilirubin remained an independent predictor. Other investigations have found total bilirubin >1.2 mg/dL significant in univariable analysis of IPMNs, implying that bilirubin may also guide risk assessment.^29^

Main duct diameter >5 mm, duct caliber change with atrophy, and mural nodules showed strong PDAC associations, aligning with established high-risk criteria.^10,11^ After adjusting for demographics and labs, main duct diameter lost significance, whereas change in duct caliber with atrophy emerged as an independent predictor. Mural nodules remained significant across all models. Cyst size showed an inverse association with PDAC in multivariable models, in contrast to evidence that larger cysts have greater malignant potential. This discrepancy may result from selection bias in a retrospective design, as larger cysts are more likely to be biopsied, leaving smaller lesions underrepresented in our study.

There are other limitations to this study. Although our NLP tools performed well within our health system, external validation is needed to assess generalizability. Moreover, performance ultimately depends on the radiologist; if an imaging feature is not documented, the LLM cannot identify it. The second phase yielded 330 patients, with only 10 low-risk cysts progressing to malignancy, restricting power in that subgroup. A prospective study is underway within our health system to actively identify PCLs, facilitate specialist referral, and encourage earlier interventions.

LLM-based entity extraction achieves outcomes comparable to manual review for PCL identification and risk stratification. Retrospective evaluation revealed factors predicting malignant progression, including older age, lower BMI, duct caliber change with upstream atrophy, mural nodules, and total bilirubin. AI-driven tools could enhance surveillance and expedite interventions for high-risk PCLs. Further multi-institutional investigations are needed to confirm generalizability.

## Data Availability

All data produced in the present study are available upon reasonable request to the authors.

## Acknowledgements

We thank the Innovation team (Northwell Health, New Hyde Park, New York) for developing and maintaining the Northwell AI Hub platform and its application programming interface. None of these individuals received financial compensation for their contributions.

