## Supplemental Tables & Figures for "Large Language Model-Based Entity Extraction Reliably Classifies Pancreatic Cysts and Reveals Predictors of Malignancy: A Cross-Sectional and Retrospective Cohort Study"

**Supplemental Online Content**

**Supplemental Table 1.** Prompts of opLLM Variants 1-3.

**Supplemental Table 2.** Distribution of pancreatic abnormalities among 14,574 radiology reports.

**Supplemental Table 3.** Patient race counts by biopsy outcome in the full cyst cohort and by subgroup.

**Supplemental Table 4.** Univariable logistic regression of pancreatic imaging features and PDAC across all cysts and subgroups.

**Supplemental Table 5.** Counts of imaging features by biopsy outcome in the entire cyst cohort and by subgroup.

**Supplemental Table 6.** Univariable logistic regression for serum laboratory studies at cyst identification by entire cohort and subgroups.

**Supplemental Table 7.** Counts and Mann-Whitney analysis of labs at cyst identification by entire cohort and subgroups.

**Supplemental Figure 1.** eeLLM Workflow for Automated Identification and Risk Stratification of Pancreatic Cysts.

**Supplemental Figure 2.** Receiver operating characteristic curves for three multivariable logistic regression models.

**Supplemental Figure 3.** Calibration plot comparing observed and predicted probabilities for three multivariable logistic regression models.

This supplemental material has been provided by the authors to give readers additional information about their work.

**Supplemental Table 1.** **Prompts of opLLM Variants 1-3.**

|  | Prompt Text |
| --- | --- |
| opLLM Variant 1 | You are a clinical researcher specialized in pancreatic diseases, tasked with reviewing CT and MRI imaging reports. Your goal is to analyze the following radiology report for specific pancreatic features. Read the report and provide your responses to the following tasks:   1. Does the report state that a pancreatic cyst is present, or is likely present? Answer “Yes” or “No”. 2. If a pancreatic cyst is present, and given the findings of the report, is this a high-risk or worrisome cyst that should be evaluated with endoscopic ultrasound and potentially fine-needle aspiration based on American College of Gastroenterology (ACG) Clinical Guidelines? Answer “Yes” or “No”. |
| opLLM Variant 2 | You are a clinical researcher specialized in pancreatic diseases, tasked with reviewing CT and MRI imaging reports. Your goal is to analyze the following radiology report for specific pancreatic features. Read the report and provide your response to the following task:   1. If a pancreatic cyst is present, and given the findings of the report, is this a high-risk or worrisome cyst that should be evaluated with endoscopic ultrasound and potentially fine-needle aspiration based on American College of Gastroenterology (ACG) Clinical Guidelines? Answer “Yes” or “No”. |
| opLLM Variant 3 | You are a clinical researcher specialized in pancreatic diseases, tasked with reviewing CT and MRI imaging reports. Your goal is to analyze the following radiology report for specific pancreatic features. Read the report and provide your response to the following task:   1. If a pancreatic cyst is present within the radiology report text provided, is this a high-risk or worrisome cyst that should be evaluated with endoscopic ultrasound and potentially fine-needle aspiration? Refer to current American College of Gastroenterology (ACG) Clinical Guidelines to help make this decision, noting that features of a high risk or worrisome cyst include: cyst size equal to or more than 3cm, cyst size equal to or more than 2cm that is not described as a possible intraductal papillary mucinous neoplasm (IPMN), pancreatic ductal dilation greater than 5mm, change in pancreatic duct caliber with concurrent pancreatic atrophy, or presence of mural nodule or solid component. Answer “Yes” or “No”. |

*Abbreviations*: opLLM, Open-Prompt Large Language Model.

**Supplemental Table 2. Distribution of pancreatic abnormalities among 14,574 radiology reports.**

|  | Count | % of Total Reports (N=14,574) | % of Abnormal Reports (N=2,301) |
| --- | --- | --- | --- |
| Any Pancreatic Abnormality | 2,301 | 15.8% | — |
| All Cysts | 665 | 4.6% | 28.9% |
| High Risk Cysts | 175 | 1.2% | 7.6% |
| Low Risk Cysts | 490 | 3.4% | 21.3% |
| Other Abnormalities | 1,636 | 11.2% | 71.1% |
| No Pancreatic Abnormality | 12,273 | 84.2% | — |

**Supplemental Table 3. Patient race counts by biopsy outcome in the full cyst cohort and by subgroup.**

|  | All Cysts | | High-Risk Cysts | | Low-Risk Cysts | |
| --- | --- | --- | --- | --- | --- | --- |
| Race | **Positive Biopsy (n=35)** | **Negative Biopsy (n=295)** | **Positive Biopsy (n=25)** | **Negative Biopsy (n=200)** | **Positive Biopsy (n=10)** | **Negative Biopsy (n=95)** |
| White | 19 | 170 | 12 | 111 | 7 | 60 |
| Black | 5 | 40 | 4 | 27 | 1 | 13 |
| Hispanic | 5 | 45 | 4 | 31 | 1 | 13 |
| Asian/Pacific Islander | 2 | 21 | 2 | 14 | 0 | 7 |
| Other | 2 | 16 | 1 | 14 | 1 | 2 |
| Unknown | 2 | 3 | 2 | 3 | 0 | 0 |

**Supplemental Table 4. Univariable logistic regression of pancreatic imaging features and PDAC across all cysts and subgroups.**

|  | All Cysts | | High-Risk Cysts | | Low-Risk Cysts | |
| --- | --- | --- | --- | --- | --- | --- |
| Imaging Features | **Odds Ratio**  **(95% CI)** | ***P*-value** | **Odds Ratio**  **(95% CI)** | ***P*-value** | **Odds Ratio**  **(95% CI)** | ***P*-value** |
| Cyst Size | 0.51  (0.25-1.03) | 0.070 | 0.20  (0.08-0.51) | 0.001* | — | — |
| Main Duct Diameter | 3.67  (1.78-7.60) | 0.001* | 4.69  (1.96-11.23) | <0.001* | — | — |
| Duct Caliber with Atrophy | 3.91  (1.65-9.29) | 0.004* | 4.12  (1.64-10.36) | 0.004* | — | — |
| Mural Nodule | 4.75  (1.52-14.82) | 0.014* | 4.75  (1.48-15.28) | 0.016* | — | — |
| Enhancing Cyst Wall | 0.48  (0.06-3.73) | 0.705 | 0.65  (0.08-5.24) | 1.000 | 0.00  (0.05-17.71) | 1.000 |
| Cyst Septations | 0.59  (0.27-1.31) | 0.263 | 0.78  (0.33-1.84) | 0.669 | 0.00  (0.01-2.49) | 0.113 |
| Pancreatitis | 0.97  (0.38-2.45) | 1.000 | 0.86  (0.31-2.42) | 1.000 | 1.40  (0.15-12.67) | 0.564 |

* signifies statistical significance.

**Supplemental Table 5. Counts of imaging features by biopsy outcome in the entire cyst cohort and by subgroup.**

|  | All Cysts | | | | High-Risk Cysts | | | | Low-Risk Cysts | | | |
| --- | --- | --- | --- | --- | --- | --- | --- | --- | --- | --- | --- | --- |
|  | **Positive Biopsy (n=35)^a^** | | **Negative Biopsy (n=295)** | | **Positive Biopsy (n=25)** | | **Negative Biopsy (n=200)** | | **Positive Biopsy (n=10)** | | **Negative Biopsy (n=95)** | |
| Imaging Features | **+** | - | **+** | - | **+** | - | **+** | - | **+** | - | **+** | - |
| Cyst Size | 15 | 20 | 176 | 119 | 15 | 10 | 176 | 24 | 0 | 10 | 0 | 95 |
| Main Duct Diameter | 16 | 19 | 55 | 240 | 16 | 9 | 55 | 145 | 0 | 10 | 0 | 95 |
| Mural Nodule | 5 | 30 | 10 | 285 | 5 | 20 | 10 | 190 | 0 | 10 | 0 | 95 |
| Duct Caliber with Atrophy | 9 | 26 | 24 | 271 | 9 | 16 | 24 | 176 | 0 | 10 | 0 | 95 |
| Enhancing Cyst Wall | 1 | 34 | 17 | 278 | 1 | 24 | 12 | 188 | 0 | 10 | 5 | 90 |
| Cyst Septations | 9 | 26 | 109 | 186 | 9 | 16 | 84 | 116 | 0 | 10 | 25 | 70 |
| Pancreatitis | 6 | 29 | 52 | 243 | 5 | 20 | 45 | 155 | 1 | 9 | 7 | 88 |

^a^ “+” indicates the feature is present and “-” indicates it is absent.

**Supplemental Table 6. Univariable logistic regression for serum laboratory studies at cyst identification by entire cohort and subgroups.**

|  | All Cysts | | High-Risk Cysts | | Low-Risk Cysts | |
| --- | --- | --- | --- | --- | --- | --- |
| Lab^a^ | **Odds Ratio (95% CI)** | ***P*-value** | **Odds Ratio (95% CI)** | ***P*-value** | **Odds Ratio (95% CI)** | ***P*-value** |
| Hemoglobin (g/dL) | 1.05  (0.83-1.32) | 0.682 | 1.02  (0.79-1.32) | 0.856 | 1.15  (0.68-1.94) | 0.596 |
| WBC Count  (K/µL) † | 0.92  (0.81-1.04) | 0.175 | 0.91  (0.79-1.05) | 0.199 | 0.88  (0.64-1.22) | 0.442 |
| BUN (mg/dL) † | 1.21  (0.51-2.86) | 0.670 | 1.58  (0.60-4.15) | 0.353 | 0.49  (0.09-2.75) | 0.414 |
| Creatinine (mg/dL) † | 0.61  (0.12-3.04) | 0.548 | 0.62  (0.11-3.61) | 0.597 | 0.46  (0.01-24.07) | 0.701 |
| Total Bilirubin (mg/dL) † | 2.12  (0.68-6.58) | 0.196 | 2.10  (0.59-7.47) | 0.251 | 2.06  (0.17-25.50) | 0.575 |
| Direct Bilirubin (mg/dL) † | 0.89  (0.20-3.87) | 0.877 | 1.14  (0.21-6.05) | 0.879 | 0.35  (0.00-55.23) | 0.687 |
| AST (U/L) † | 1.03  (0.58-1.82) | 0.921 | 0.74  (0.34-1.62) | 0.447 | 2.02  (0.79-5.20) | 0.143 |
| ALT (U/L) † | 0.89  (0.52-1.53) | 0.666 | 0.64  (0.30-1.35) | 0.243 | 1.59  (0.68-3.71) | 0.282 |
| ALP (U/L) † | 0.93  (0.36-2.40) | 0.886 | 0.65  (0.20-2.10) | 0.469 | 2.00  (0.42-9.49) | 0.382 |
| Albumin (g/dL) | 0.97  (0.47-2.02) | 0.940 | 1.27  (0.52-3.10) | 0.603 | 0.54  (0.15-2.00) | 0.356 |
| Lipase  (U/L) † | 0.98  (0.66-1.46) | 0.918 | 0.68  (0.38-1.22) | 0.194 | 2.37  (0.96-5.87) | 0.062 |
| CRP  (mg/L) † | 1.25  (0.62-2.53) | 0.534 | 1.19  (0.36-3.89) | 0.780 | 1.37  (0.55-3.44) | 0.503 |
| ESR (mm/h) | 1.00  (0.97-1.04) | 0.801 | 1.03  (0.98-1.08) | 0.257 | 0.98  (0.92-1.04) | 0.441 |
| HbA1c  (%) | 1.15  (0.81-1.64) | 0.445 | 1.30  (0.89-1.91) | 0.180 | 0.63  (0.21-1.94) | 0.423 |
| CA 19-9  (U/mL) † | 3.42  (1.24-9.45) | 0.018* | 3.20  (1.18-8.67) | 0.023* | — | — |
| CEA  (ng/mL) † | 1.72  (0.56-5.26) | 0.340 | 9.26  (0.59-145.94) | 0.114 | — | — |
| AFP (ng/mL) | 3.26  (0.66-16.18) | 0.148 | 2.89  (0.63-13.22) | 0.171 | — | — |

* signifies statistical significance.

^a^ “†” marker indicates that the lab was log-transformed in the regression model to address skewness.

*Abbreviations:* AFP, alpha-fetoprotein; ALP, alkaline phosphatase; ALT, alanine aminotransferase; AST, aspartate aminotransferase; BUN, blood urea nitrogen; CA 19-9, carbohydrate antigen 19-9; CEA, carcinoembryonic antigen; CRP, C-reactive protein; ESR, erythrocyte sedimentation rate; HbA1C, hemoglobin A1c; WBC, white blood cell.

**Supplemental Table 7. Counts and Mann-Whitney analysis of labs at cyst identification by entire cohort and subgroups.**

| A. All Cysts | | | | | | |
| --- | --- | --- | --- | --- | --- | --- |
|  | **Counts** | | | **Mann-Whitney Analysis** | | |
| Lab | **Total** | **PDAC-positive** | **PDAC-negative** | **PDAC-positive**  **Median (IQR)** | **PDAC-negative**  **Median (IQR)** | ***P*-value** |
| Hemoglobin (g/dL) | 267 | 25 | 242 | 13.4 (2.2) | 13.4 (2.28) | 0.758 |
| WBC (×10^3/µL) | 267 | 25 | 242 | 6.55 (3.09) | 8.22 (4.59) | 0.221 |
| BUN (mg/dL) | 271 | 26 | 245 | 18.0 (12.5) | 16.0 (10.0) | 0.585 |
| Creatinine (mg/dL) | 272 | 26 | 246 | 0.8 (0.34) | 0.93 (0.43) | 0.528 |
| Total Bilirubin (mg/dL) | 261 | 26 | 235 | 0.5 (0.48) | 0.5 (0.4) | 0.236 |
| Direct Bilirubin (mg/dL) | 70 | 7 | 63 | 0.2 (0.2) | 0.2 (0.4) | 0.708 |
| AST (U/L) | 264 | 26 | 238 | 23.0 (16.75) | 22.0 (15.75) | 0.723 |
| ALT (U/L) | 264 | 26 | 238 | 21.5 (27.25) | 21.5 (18.0) | 0.839 |
| ALP (U/L) | 264 | 26 | 238 | 76.5 (33.0) | 77.5 (37.0) | 0.599 |
| Albumin (g/dL) | 264 | 26 | 238 | 4.05 (0.65) | 4.2 (0.7) | 0.657 |
| Lipase (U/L) | 139 | 10 | 129 | 111.0 (86.25) | 100.0 (211.0) | 0.880 |
| CRP (mg/L) | 44 | 3 | 41 | 17.13 (39.25) | 9.0 (37.1) | 0.530 |
| ESR (mm/h) | 42 | 4 | 38 | 43.5 (26.5) | 33.5 (44.25) | 0.748 |
| HbA1c  (%) | 116 | 14 | 102 | 5.9 (1.55) | 5.9 (1.07) | 1.000 |
| CA 19-9 (U/mL) | 54 | 6 | 48 | 70.85 (53.07) | 12.0 (18.25) | 0.006* |
| CEA (ng/mL) | 42 | 3 | 39 | 4.2 (3.6) | 1.9 (1.45) | 0.150 |
| AFP (ng/mL) | 19 | 2 | 17 | 6.6 (1.0) | 3.6 (1.9) | 0.096 |
| B. High-Risk Cysts | | | | | | |
|  | **Counts** | | | **Mann-Whitney Analysis** | | |
| Lab | **Total** | **PDAC-positive** | **PDAC-negative** | **PDAC-positive**  **Median (IQR)** | **PDAC-negative**  **Median (IQR)** | ***P*-value** |
| Hemoglobin (g/dL) | 182 | 19 | 163 | 13.3 (2.2) | 13.4 (2.3) | 0.887 |
| WBC (×10^3/µL) | 182 | 19 | 163 | 7.2 (3.32) | 8.63 (4.59) | 0.290 |
| BUN (mg/dL) | 185 | 19 | 166 | 19.0 (12.5) | 16.0 (9.0) | 0.238 |
| Creatinine (mg/dL) | 186 | 19 | 167 | 0.85 (0.43) | 0.95 (0.43) | 0.676 |
| Total Bilirubin (mg/dL) | 178 | 19 | 159 | 0.6 (0.6) | 0.5 (0.4) | 0.272 |
| Direct Bilirubin (mg/dL) | 52 | 5 | 47 | 0.2 (0.3) | 0.3 (0.37) | 0.873 |
| AST (U/L) | 180 | 19 | 161 | 21.0 (15.0) | 22.0 (18.0) | 0.616 |
| ALT (U/L) | 180 | 19 | 161 | 20.0 (21.0) | 22.0 (19.0) | 0.326 |
| ALP (U/L) | 180 | 19 | 161 | 72.0 (28.5) | 77.0 (38.0) | 0.421 |
| Albumin (g/dL) | 180 | 19 | 161 | 4.1 (0.6) | 4.1 (0.7) | 0.749 |
| Lipase (U/L) | 103 | 8 | 95 | 89.5 (61.5) | 140.5 (370.1) | 0.225 |
| CRP (mg/L) | 33 | 1 | 32 | 17.13 (0.0) | 12.0 (34.97) | 1.000 |
| ESR (mm/h) | 30 | 2 | 28 | 57.0 (5.0) | 28.0 (38.25) | 0.170 |
| HbA1c  (%) | 80 | 10 | 70 | 6.05 (2.03) | 5.9 (1.0) | 0.802 |
| CA 19-9 (U/mL) | 44 | 6 | 38 | 70.85 (53.07) | 12.0 (17.0) | 0.009* |
| CEA (ng/mL) | 28 | 3 | 25 | 4.2 (3.6) | 2.1 (1.4) | 0.194 |
| AFP (ng/mL) | 12 | 2 | 10 | 6.6 (1.0) | 4.2 (0.7) | 0.233 |
| C. Low-Risk Cysts | | | | | | |
|  | **Counts** | | | **Mann-Whitney Analysis** | | |
| Lab | **Total** | **PDAC-positive** | **PDAC-negative** | **PDAC-positive**  **Median (IQR)** | **PDAC-negative**  **Median (IQR)** | ***P*-value** |
| Hemoglobin (g/dL) | 85 | 6 | 79 | 13.6 (1.75) | 13.5 (1.9) | 0.657 |
| WBC (×10^3/µL) | 85 | 6 | 79 | 5.88 (2.06) | 7.82 (3.14) | 0.189 |
| BUN (mg/dL) | 86 | 7 | 79 | 13.0 (8.0) | 17.0 (10.0) | 0.334 |
| Creatinine (mg/dL) | 86 | 7 | 79 | 0.77 (0.13) | 0.9 (0.4) | 0.573 |
| Total Bilirubin (mg/dL) | 83 | 7 | 76 | 0.4 (0.3) | 0.5 (0.38) | 0.625 |
| Direct Bilirubin (mg/dL) | 18 | 2 | 16 | 0.25 (0.05) | 0.2 (0.2) | 0.590 |
| AST (U/L) | 84 | 7 | 77 | 33.0 (43.0) | 21.0 (14.0) | 0.169 |
| ALT (U/L) | 84 | 7 | 77 | 22.0 (47.0) | 21.0 (18.0) | 0.255 |
| ALP (U/L) | 84 | 7 | 77 | 87.0 (43.0) | 78.0 (37.5) | 0.794 |
| Albumin (g/dL) | 84 | 7 | 77 | 4.0 (0.5) | 4.2 (0.6) | 0.223 |
| Lipase (U/L) | 36 | 2 | 34 | 211.0 (52.0) | 46.0 (81.0) | 0.100 |
| CRP (mg/L) | 11 | 2 | 9 | 43.25 (39.25) | 3.43 (44.78) | 0.389 |
| ESR (mm/h) | 12 | 2 | 10 | 21.0 (14.0) | 44.0 (59.0) | 0.829 |
| HbA1c  (%) | 36 | 4 | 32 | 5.85 (0.60) | 6.0 (1.9) | 0.642 |
| CA 19-9 (U/mL) | 10 | 0 | 10 | — | — | — |
| CEA (ng/mL) | 14 | 0 | 14 | — | — | — |
| AFP (ng/mL) | 7 | 0 | 7 | — | — | — |

* signifies statistical significance.

*Abbreviations:* AFP, alpha-fetoprotein; ALP, alkaline phosphatase; ALT, alanine aminotransferase; AST, aspartate aminotransferase; BUN, blood urea nitrogen; CA 19-9, carbohydrate antigen 19-9; CEA, carcinoembryonic antigen; CRP, C-reactive protein; ESR, erythrocyte sedimentation rate; HbA1c, hemoglobin A1c; IQR, interquartile range; PDAC, pancreatic ductal adenocarcinoma; WBC, white blood cell.

**Supplemental Figure 1. eeLLM Workflow for Automated Identification and Risk Stratification of Pancreatic Cysts.**

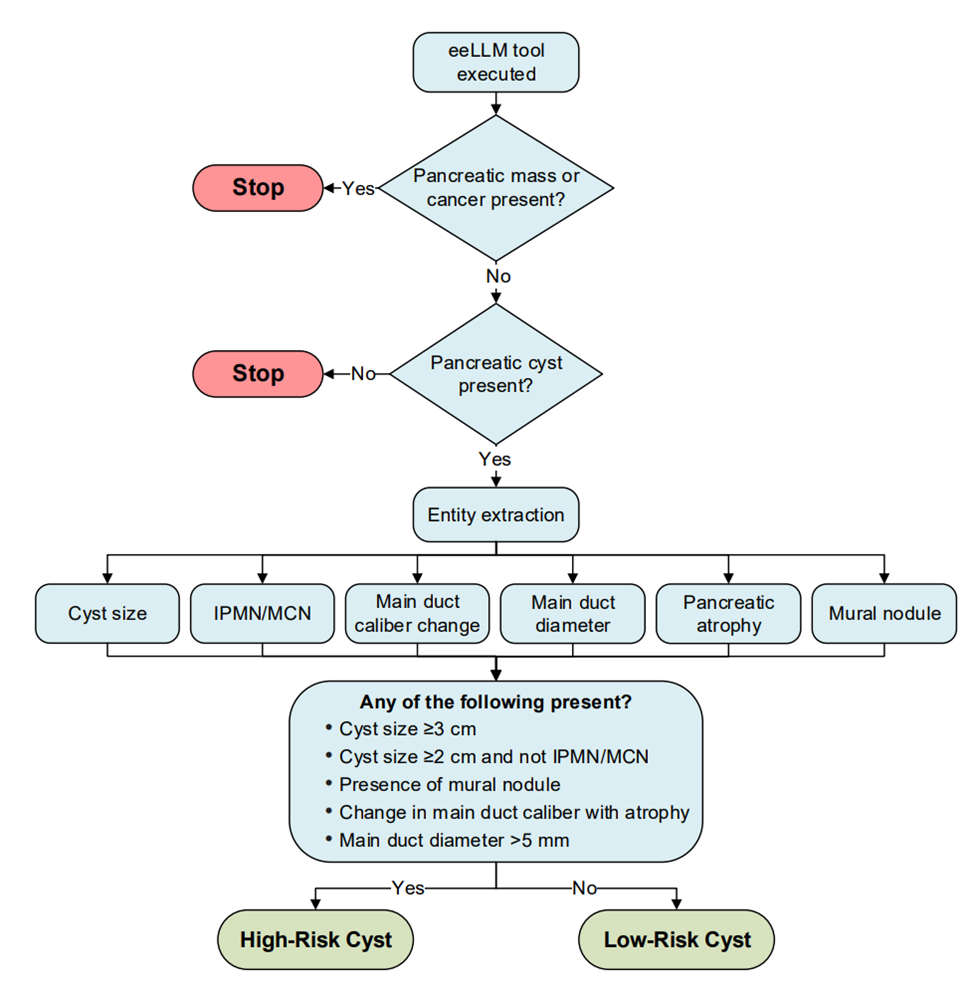

*Abbreviations*: eeLLM, Entity-Extraction Large Language Model; IPMN, intraductal papillary mucinous neoplasm; MCN, mucinous cystic neoplasm.

**Supplemental Figure 2. Receiver operating characteristic curves for three multivariable logistic regression models.**

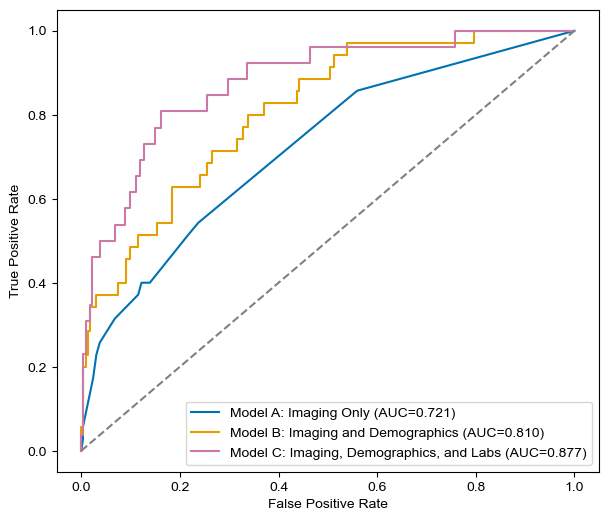

*Abbreviations*: AUC, area under the curve.

**Supplemental Figure 3. Calibration plot comparing observed and predicted probabilities for three multivariable logistic regression models.**

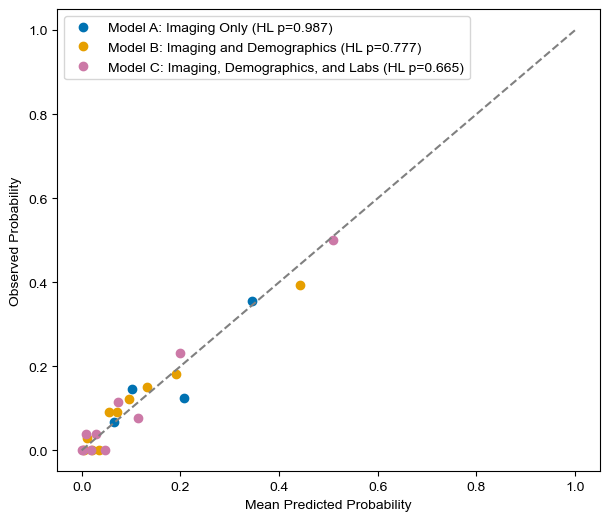

HL *P*>0.05 indicates no significant deviation from perfect calibration. *Abbreviations:* HL, Hosmer-Lemeshow.
